# Post-peak dynamics of a national Omicron SARS-CoV-2 epidemic during January 2022

**DOI:** 10.1101/2022.02.03.22270365

**Authors:** Paul Elliott, Oliver Eales, Barbara Bodinier, David Tang, Haowei Wang, Jakob Jonnerby, David Haw, Joshua Elliott, Matthew Whitaker, Caroline E. Walters, Christina Atchison, Peter J. Diggle, Andrew J. Page, Alexander J. Trotter, Deborah Ashby, Wendy Barclay, Graham Taylor, Helen Ward, Ara Darzi, Graham S. Cooke, Marc Chadeau-Hyam, Christl A. Donnelly

## Abstract

**Background:** Rapid transmission of the SARS-CoV-2 Omicron variant has led to the highest ever recorded case incidence levels in many countries around the world.

**Methods:** The REal-time Assessment of Community Transmission-1 (REACT-1) study has been characterising the transmission of the SARS-CoV-2 virus using RT-PCR test results from self-administered throat and nose swabs from randomly-selected participants in England at ages 5 years and over, approximately monthly since May 2020. Round 17 data were collected between 5 and 20 January 2022 and provide data on the temporal, socio-demographic and geographical spread of the virus, viral loads and viral genome sequence data for positive swabs.

**Results:** From 102,174 valid tests in round 17, weighted prevalence of swab positivity was 4.41% (95% credible interval [CrI], 4.25% to 4.56%), which is over three-fold higher than in December 2021 in England. Of 3,028 sequenced positive swabs, 2,393 lineages were determined and 2,374 (99.2%) were Omicron including 19 (0.80% of all Omicron lineages) cases of BA.2 sub-lineage and one BA.3 (0.04% of all Omicron) detected on 17 January 2022, and only 19 (0.79%) were Delta. The growth of the BA.2 Omicron sub-lineage against BA.1 and its sub-lineage BA.1.1 indicated a daily growth rate advantage of 0.14 (95% CrI, 0.03, 0.28) for BA.2, which corresponds to an additive R advantage of 0.46 (95% CrI, 0.10, 0.92).

Within round 17, prevalence was decreasing overall (R=0.95, 95% CrI, 0.93, 0.97) but increasing in children aged 5 to 17 years (R=1.13, 95% CrI, 1.09, 1.18). Those 75 years and older had a swab-positivity prevalence of 2.46% (95% CI, 2.16%, 2.80%) reflecting a high level of infection among a highly vulnerable group. Among the 3,613 swab-positive individuals reporting whether or not they had had previous infection, 2,334 (64.6%) reported previous confirmed COVID-19. Of these, 64.4% reported a positive test from 1 to 30 days before their swab date. Risks of infection were increased among essential/key workers (other than healthcare or care home workers) with mutually adjusted Odds Ratio (OR) of 1.15 (95% CI, 1.05, 1.26), people living in large compared to single-person households (6+ household size OR 1.73; 95% CI, 1.44, 2.08), those living in urban vs rural areas (OR 1.24, 95% CI, 1.13, 1.35) and those living in the most vs least deprived areas (OR 1.34, 95% CI, 1.20, 1.49).

**Conclusions:** We observed unprecedented levels of infection with SARS-CoV-2 in England in January 2022, an almost complete replacement of Delta by Omicron, and evidence for a growth advantage for BA.2 compared to BA.1. The increase in the prevalence of infection with Omicron among children (aged 5 to 17 years) during January 2022 could pose a risk to adults, despite the current trend for prevalence in adults to decline. (Funded by the Department of Health and Social Care in England.)

## Introduction

November 2021 saw the identification of the Omicron variant in Botswana and South Africa, its rapid replacement of the Delta (B.1.617.2) variant within South Africa,^1^ and on 26 November the classification by WHO of Omicron as a variant of concern.^2^ By 1 December 2021, Omicron had been identified in the UK^3^ and the USA^4^ as well as other countries including Belgium, Hong Kong and Israel, initially in travel-related cases.

By mid-to late December 2021, Omicron had become the dominant variant in the UK^5,6^ and had been detected in most European countries and US states. Not only was the increase in Omicron in England^5^ and elsewhere^7^ extremely rapid, but replacement of Omicron by Delta was over three-times faster than that of Alpha by Delta^5^. In some countries, social distancing policies were brought back into force^8^ and vaccination programmes accelerated,^9^ while some health care systems struggled to cope with the associated increased health care demands.^10,11^ The REal-time Assessment of Community Transmission-1 (REACT-1) study has been tracking the spread of the SARS-CoV-2 virus among randomly-selected community samples in England, approximately monthly since May 2020, avoiding the biases associated with case incidence data and the delays inherent in hospitalizations and deaths.^12^ We use recent rounds of the REACT-1 study to document the transmission dynamics of SARS-CoV-2 in England with a particular focus on Omicron (including the BA.2 and BA.3 sub-lineages) during January 2022.

## Methods

The REACT-1 study involves a series of cross-sectional surveys of random samples of the population of England at ages 5 years and over,^13^conducted approximately monthly over a two-to three-week period since May 2020 (exceptions were December 2020 and August 2021). The present report is for round 17 (5 to 20 January 2022) involving N=102,174 participants with a valid self-administered throat and nose swab test result for SARS-CoV-2 by reverse transcription polymerase chain reaction (RT-PCR) (including 862 samples [36 positives] obtained between 21 and 24 January 2022). A positive test result was recorded if both N gene and E gene targets were detected or if N gene was detected with cycle threshold (Ct) value below 37. We also tested for influenza A and B using multiplex PCR. We compare results for SARS-CoV-2 with those obtained during round 15 (19 October to 5 November 2021, N=100,112, including 93 samples from 6-8 November)^14^ and round 16 (23 November to 14 December 2021, N=97,089, including 661 samples from 15-17 December 2021).^5^ We used as the sampling frame the general practitioner list of patients in England held by the National Health Service (NHS). Participants completed a brief registration and an online or telephone questionnaire.^15^ We obtained information on age, sex, residential postcode, ethnicity, household size, occupation, potential contact with a COVID-19 case, symptoms and other variables. We used the postcode of residence to link to an area-level Index of Multiple Deprivation^16^ and urban/rural status.^17^

Initially we aimed to obtain approximately equal numbers of participants in each lower-tier local authority (LTLA) in England (N=315), but from round 12 (20 May to 7 June 2021) we switched to obtaining a random sample in proportion to population size at LTLA level. We use random iterative method (rim) weighting^18^ to provide prevalence estimates for the population of England as a whole, adjusting for age, sex, deciles of the Index of Multiple Deprivation, LTLA counts, and ethnic group. Up to round 13 (24 June to 12 July 2021), we collected dry swabs sent by courier to the laboratory on a cold chain but from round 14 (9 to 27 September 2021 including 509 samples from 28-30 September) we switched to ‘wet’ (saline) swabs which (round 14) were sent to the laboratory either by courier (no cold chain) or priority post, and from round 15 onwards by priority post only. Because of delays in the post for return of swabs, we include a small proportion of samples obtained after the nominated closing date for the study.

Samples testing positive with Ct 34 or less in either the E or N gene were sent for viral genome sequencing to the Quadram Institute, Norwich, UK. RT-PCR was performed on 96 randomly chosen samples using the CDC assay^19^ by the Quadram Institute as a secondary confirmation of the Ct values. We used the ARTIC protocol^20^ (version 4) for viral RNA amplification, CoronaHiT for preparation of sequencing libraries,^21^ the ARTIC bioinformatics pipeline^20^ and assigned lineages using PangoLEARN (version 2022-01-20).^22^

### Data analyses

We estimated weighted prevalence and 95% credible intervals overall and by socio-demographic and other variables, comparing round 17 to round 16. We used logistic regression to estimate the odds of testing positive by employment, ethnicity, household size, children in household, urban area, and deprivation, adjusting for age, region and the other variables examined.

We fit a Bayesian logistic regression model to the proportion of BA.2 lineage compared to the BA.1 lineage (and its sub-lineage BA.1.1) during round 17 to investigate whether there was a daily growth rate advantage for the odds of BA.2 versus BA.1. The daily percentage growth in the odds of BA.2 infection was estimated from the exponential of the daily growth rate. The estimated additive R advantage was estimated as the daily growth rate advantage multiplied by the Omicron-specific mean generation time.^23^

We used an exponential model of growth or decay to examine temporal trends in swab positivity assuming a binomial distribution for the numbers of positives out of the total number of samples per day. To estimate the growth rate and posterior credible intervals, we used day of sampling where reported (otherwise day of first scan of the swab by the Post Office if available) with a bivariate No-U-Turn Sampler and uniform prior distribution for the probability of swab positivity.^24^ For the reproduction number R, we assumed a gamma distribution in the generation time with Omicron-specific mean 3.3 days and standard deviation 3.5 days, setting the shape parameter n to 0.89 and rate parameter *β*to 0.27:^23^

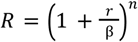

where *r* is the exponential growth (or decay) rate.

To visualise temporal trends in swab positivity, we used a No-U-Turn Sampler in logit space to fit a Bayesian penalised-spline (P-spline) model^25^ to the daily data, split into approximately 5-day sections by regularly spaced knots. Edge effects were minimised by adding further knots beyond the study period. We used fourth-order basis splines (b-splines) over the knots including a second-order random-walk prior distribution on the coefficients of the b-splines to guard against overfitting; the prior penalised against changes in growth rate unless supported by the data.^26^ We also fit P-splines separately to three broad age groups (17 years and under, 18 to 54 years, 55 years and over) with smoothing parameter obtained from the model fit to all the data.

We compared Ct values (using a Kruskal–Wallis test) among test-positive swabs (N gene and E gene where Ct>0) as a proxy for viral load, by vaccination status (lagged by a 14-day period from date of vaccination) and symptom status across rounds 15 to 17, where information on vaccination history and dates of vaccination was obtained (with consent) by linking to data from the national COVID-19 vaccination programme.

We used a neighbourhood spatial smoothing method to examine geographical variation in SARS-CoV-2 prevalence at the LTLA level. For each of 15 randomly selected participants within an LTLA, we calculated the prevalence of infection among the nearest M people, where M was the median number of study participants within 30 km, and then estimated the smoothed neighbourhood prevalence in that area.

We then compared swab positivity prevalence in REACT-1 with daily hospitalisations and (separately) deaths from (external) national data. Estimates of the lag and scaling parameters to fit these different datasets were obtained using REACT-1 data up to round 7 (13 November to 3 December 2020), before the vaccination programme in England began. To account for variant-specific lags, mostly reflecting the dynamics of disease progression, we estimated a second lag parameter from round 13 (24 June to 12 July 2021), when Delta became dominant in England, to round 16 (23 November to 14 December 2021^1^). We used the original estimated scaling factor throughout with the original lag estimate for round 1 (01 May to 01 June 2020) to round 12 (20 May to 07 June 2021), and the second lag estimate thereafter.

We used R software^27^ for the data analyses.

## Results

Of the 840,530 invited participants, 140,075 (16.7%) registered and 102,279 (72.9%) returned a self-administered throat and nasal swab to date, of whom 102,174 (99.9%) provided a valid SARS-CoV-2 RT-PCR test result (Supplementary Figure 1). Of these 102,174 valid tests, 4,073 were positive yielding a weighted prevalence of 4.41% (95% credible interval [CrI], 4.25%, 4.56%), which is over three-fold higher than the weighted prevalence of round 16 at 1.40% (95% CrI, 1.31%, 1.50%) (Supplementary Table 1).

A total of 3,028 positive swab samples were sequenced resulting in 2,393 (79.0%, 95% CI, 77.5%, 80.5%) determined viral genomes (Supplementary Table 2). Of these 2,374 (99.2%, 95% CI, 98.8%, 99.5%) were Omicron and 19 (0.79%, 95% CI, 0.48%, 1.24%) Delta or Delta sub-lineages. Of the 2,374 Omicron lineages 13 (0.55%, 95% CI, 0.29%, 0.93%) were B.1.1.529, 1,822 (76.8%, 95% CI, 75.0%, 78.4%) were BA.1, 519 (21.9%, 95% CI 20.2%, 23.6%) were BA.1.1, 19 (0.80%, 95% CI, 0.48%, 1.25%) were BA.2, and 1 (0.04% 95% CI 0.00%, 0.23%) was BA.3, detected on 17 January 2022 in South East. Exponential models indicated a daily growth of 1.14 (95% CrI 1.03, 1.28) in the odds of BA.2 (vs BA.1 or BA.1.1), with the proportion of BA.2 estimated at 2.84% (95% CrI, 0.97%, 6.98%) on 20 January 2022 (Supplementary Figure 2). The growth rate advantage (0.14, 95% CrI 0.03, 0.28) corresponds to an additive R advantage of 0.46 (95% CrI, 0.10, 0.92).

Using a P-spline constrained by parameters estimated for the whole period of REACT-1, we observed a substantial increase in weighted prevalence between round 16 (23 November to 14 December 2021)^2^ and round 17 (5 to 20 January 2022)^3^ (Figure 1A). Within-round estimated reproduction number was 0.95 (95% CrI, 0.93, 0.97) with less than 0.01 posterior probability that R>1 (Table 1).

**Table 1.** Table of growth rates per day (r), reproduction numbers (R) and doubling/halving times (in days) of SARS-CoV-2 swab-positivity from exponential model fits on data from round 17 (05 to 20 January 2022)^4^

| Rounds |  |  | Growth rate per day (r) | Reproduction number (R)* | Probability R>1, r>0 | Doubling (+) / Halving (-) time (in days) |
| --- | --- | --- | --- | --- | --- | --- |
| 17 | All positives |  | -0.015 ( -0.022 , -0.008 ) | 0.95 ( 0.93 , 0.97 ) | <0.01 | -46.7 ( -31.2 , -90.8 ) |
|  | Age | Aged 17 and under | 0.040 ( 0.026 , 0.054 ) | 1.13 ( 1.09 , 1.18 ) | >0.99 | 17.3 ( 26.5 , 12.9 ) |
|  |  | Aged 18 to 54 | -0.045 ( -0.055 , -0.035 ) | 0.85 ( 0.82 , 0.89 ) | <0.01 | -15.5 ( -12.6 , -20.1 ) |
|  |  | Aged 55 and over | -0.032 ( -0.047 , -0.016 ) | 0.89 ( 0.84 , 0.95 ) | <0.01 | -21.7 ( -14.6 , -42.1 ) |
|  | Region | East Midlands | -0.009 ( -0.035 , 0.017 ) | 0.97 ( 0.88 , 1.06 ) | 0.26 | -79.7 ( -19.7 , 40.3 ) |
|  |  | West Midlands | 0.004 ( -0.016 , 0.024 ) | 1.01 ( 0.95 , 1.08 ) | 0.66 | 161.2 ( -43.1 , 28.4 ) |
|  |  | East of England | -0.008 ( -0.033 , 0.016 ) | 0.97 ( 0.89 , 1.05 ) | 0.26 | -84.3 ( -21.0 , 42.4 ) |
|  |  | London | -0.014 ( -0.031 , 0.003 ) | 0.96 ( 0.90 , 1.01 ) | 0.06 | -51.1 ( -22.5 , 198.3 ) |
|  |  | North West | -0.034 ( -0.053 , -0.016 ) | 0.89 ( 0.82 , 0.95 ) | <0.01 | -20.2 ( -13.0 , -44.2 ) |
|  |  | North East | -0.008 ( -0.036 , 0.018 ) | 0.97 ( 0.88 , 1.06 ) | 0.27 | -82.9 ( -19.4 , 38.1 ) |
|  |  | South East | -0.009 ( -0.030 , 0.012 ) | 0.97 ( 0.90 , 1.04 ) | 0.21 | -79.4 ( -23.4 , 58.5 ) |
|  |  | South West | -0.003 ( -0.030 , 0.024 ) | 0.99 ( 0.90 , 1.08 ) | 0.41 | -218.4 ( -22.8 , 29.0 ) |
|  |  | Yorkshire and The Humber | -0.046 ( -0.068 , -0.025 ) | 0.85 ( 0.77 , 0.92 ) | <0.01 | -15.1 ( -10.2 , -28.1 ) |

\* Within-round R was calculated assuming an Omicron-specific Gamma-distributed generation time with mean 3.3 days and standard deviation of 3.5 days
<sup>4</sup> N=862 (including 36 positives) from 21-24 January 2022

**Figure 1.**
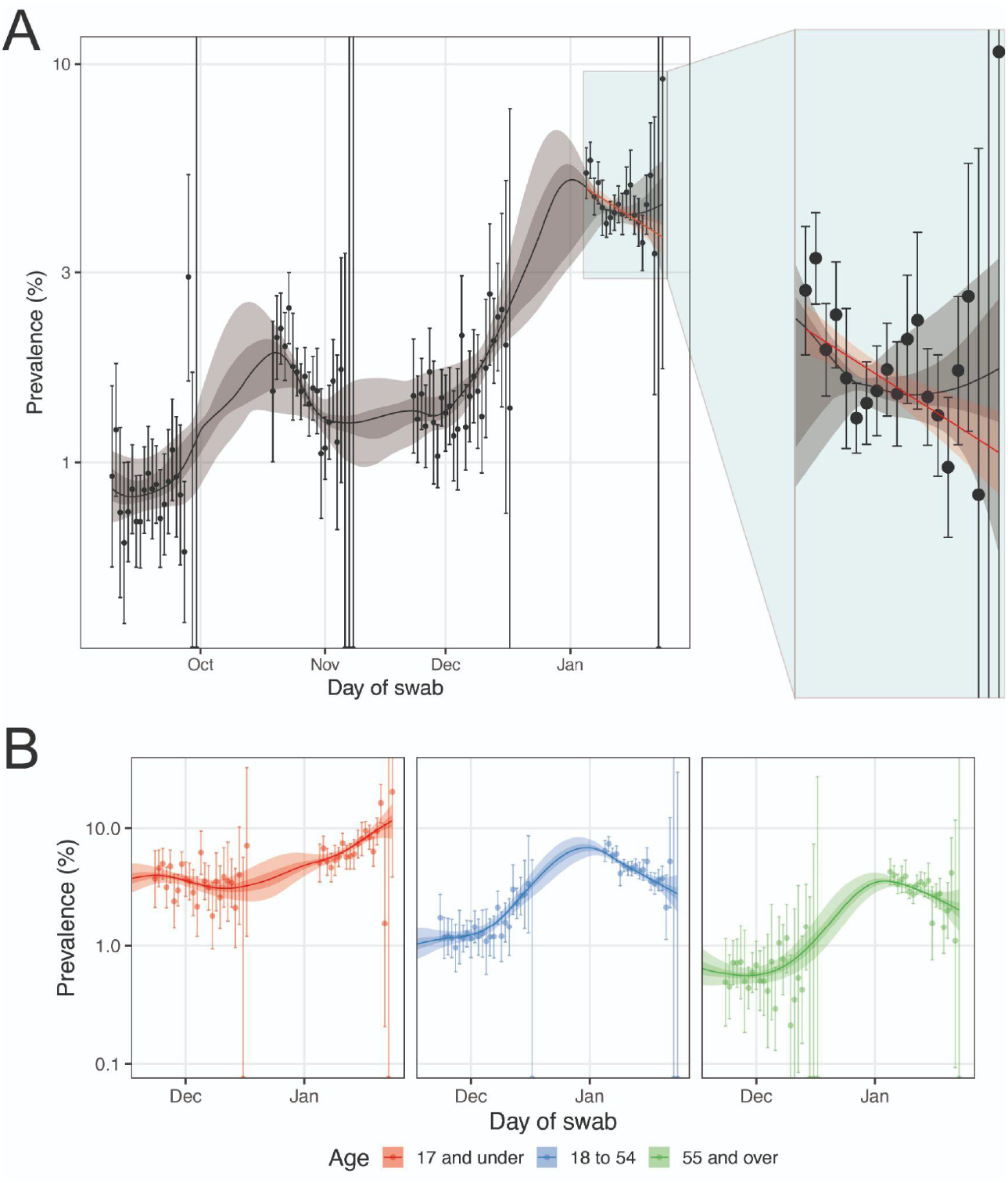
Comparison of an exponential model fit to SARS-CoV-2 swab-positivity data in round 17 (red), and a P-spline model fit to all rounds of REACT-1 (black, shown here only for rounds 14, 15, 16 and 17) (A). Shaded red region shows the 95% posterior credible interval for the exponential models, and the shaded grey region shows 50% (dark grey) and 95% (light grey) posterior credible interval for the P-spline model. Results are presented for each day (X axis) of sampling for round 14, round 15, round 16 and round 17 and the weighted prevalence of swab-positivity is shown (Y axis) on a log scale. Weighted observations (black dots) and 95% confidence intervals (vertical lines) are also shown. Results from similar P-spline models for those aged 17 years and under (red), those aged 18 to 54 years inclusive (blue) and those aged 55 years and over (green) (B). Results are presented for round 16 and 17.

P-splines stratified by age group showed a within-round 17 increasing weighted prevalence in those aged 17 years and under (Figure 1B) and corresponding within-round R of 1.13 (95% CrI, 1.09, 1.18) with greater than 0.99 posterior probability that R>1 (Table 1). In those aged 18 to 54 years R was 0.85 (95% CrI, 0.82, 0.89) and it was 0.89 (95% CrI, 0.84, 0.95) for those aged 55 years and over, both having less than 0.01 posterior probability that R>1.

Region-specific exponential models showed a decrease in weighted prevalence within round 17 in the north of the country (Yorkshire and The Humber and North West) with within-round 17 R estimates of 0.85 (95% CrI, 0.77, 0.92) and 0.89 (95% CrI, 0.82, 0.95), respectively (Table 1).

In adults who received three vs two vaccine doses, we observed higher Ct values (lower viral load) in round 16 for N and E gene (when Delta predominated), and also E gene in round 15 (all Delta), but not in round 17 (predominantly Omicron). In round 15, round 16 and round 17 we found lower Ct values (higher viral load) for N gene in swab-positives reporting symptoms one month prior to swabbing, compared to those not reporting any symptoms (p<0.01) (Figure 2). The distributions of Ct values of both E and N genes were similar in unvaccinated and vaccinated children aged 17 years and below in round 15, round 16 and round 17.

**Figure 2.**
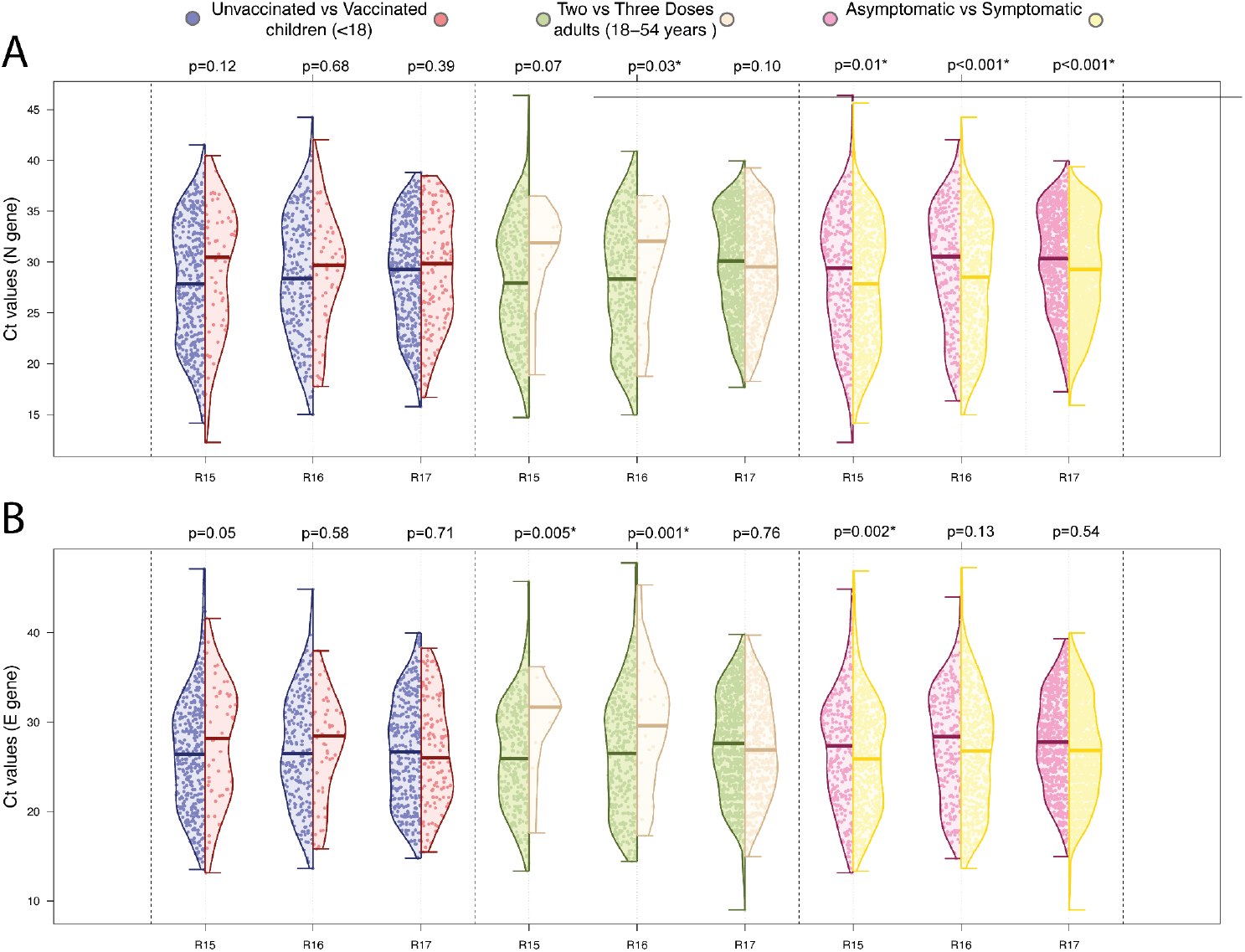
Distribution of the Ct value for the N gene (A) and E gene (B) in swab-positive samples from round 15 (Delta), round 16 (predominantly Delta) and round 17 (predominantly Omicron). Within each round, distributions are compared (i) for vaccinated vs. unvaccinated participants aged 17 years and under, (ii) those having received a three vs two vaccine doses in adults aged 18 to 54 years, and (iii) for those reporting any symptoms vs those not reporting any symptom in the month prior to swabbing. For each comparison, we report the P-value from a non-parametric (Kruskal-Wallis) test.

At all ages, weighted prevalence increased from two-(in those aged 5 to 11 years) to almost twelve-fold (in those ages 75 years and over) between round 16 and round 17 (Supplementary Table 3A, Supplementary Figure 3). The highest weighted prevalence in round 17 was observed in those aged 5 to 11 years at 7.85% (95% CrI, 7.10%, 8.69%) and the lowest in those ages 75 years and over at 2.46% (95% CrI, 2.16%, 2.80%). Weighted prevalence also increased between rounds in all regions (Supplementary Table 3A, Supplementary Figures 4 and 5), ranging in round 17 from 6.86% (95% CrI, 5.99%, 7.84%) in North East to 2.92% (95% CrI, 2.58%, 3.30%) in South East.

Our results are suggestive of within-household transmission with weighted prevalence increasing with (i) size of household from 3.15% (95% CrI, 2.86%, 3.48%) in single-person households to 7.72% (95% CrI, 6.47%, 9.18%) in households with 6 or more persons and (ii) number of children in the household from 3.57% (95% CrI, 3.41%, 3.74%) in households without children to 5.92% (95% CrI, 5.60%, 6.25%) in households with one or more children (Supplementary Table 3B).

We also found higher weighted prevalence in those having been in contact with a confirmed (12.8%, 95% CrI, 12.2%, 13.5%) or a suspected COVID-19 case (9.38%, 95% CrI, 8.14%, 10.8%) compared to 2.40% (95% CrI, 2.27%, 2.54%) for those without such contact, and in those reporting classic COVID-19 symptoms (loss or change of sense of smell or taste, fever, new persistent cough) in the month prior to swabbing at 15.9% (95% CrI, 15.1%, 16.7%) compared to 1.87% (95% CrI, 1.74%, 2.01%) in those without symptoms. Weighted prevalence in those who reported they were shielding was 3.44% (95% CrI, 3.18%, 3.73%) compared to 4.60% (95% CrI, 4.41%, 4.80%) in those not shielding.

Those who reported previous confirmed COVID-19 had a higher weighted prevalence (14.0%, 95% CrI, 13.4%, 14.6%) than those who reported previous suspected COVID-19 (3.84%, 95% CrI, 3.32%, 4.43%) or no previous COVID-19 (1.75%, 95% CrI, 1.64%, 1.88%) (Supplementary Table 3B). Among the 2,334 swab-positive individuals reporting previous confirmed COVID-19, 1,771 (75.9%) also provided a date for their most recent positive test, of which 153 were invalid (on or after the swab date); 1,337 (57.3%) reported a previous positive test 1 to 14 days, 168 (7.16%) 15 to 30 days, and 114 (4.88%) more than 30 days prior to swabbing (Supplementary Table 4). Among the 13,865 participant testing negative and reporting previous confirmed COVID-19, 10,747 (77.5%) provided a date for their most recent positive test: 904 (6.52%) were 1 to 14 days, 2,032 (14.7%) were 15 to 30 days, and 7,761 (56.0%) were more than 30 days prior to swabbing.

Multivariable logistic regression models identified variables independently associated with higher risk of swab-positivity including (i) being an essential/key worker (other than a healthcare or care home worker) with mutually adjusted Odds Ratio (OR) of 1.15 (95% CI, 1.05, 1.26), living in a large household with OR of 1.28 (95% CI 1.17, 1.40) and of 1.73 (95% CI 1.44, 2.08) for households of 3 to 5 and 6 or more persons, respectively, compared to single-person households, (iii) living in urban (vs. rural) areas with OR of 1.24 (95% CI 1.13, 1.35), and living in the most vs least deprived areas with OR of 1.34 (95% CI 1.20, 1.49) (Table 2).

**Table 2.**
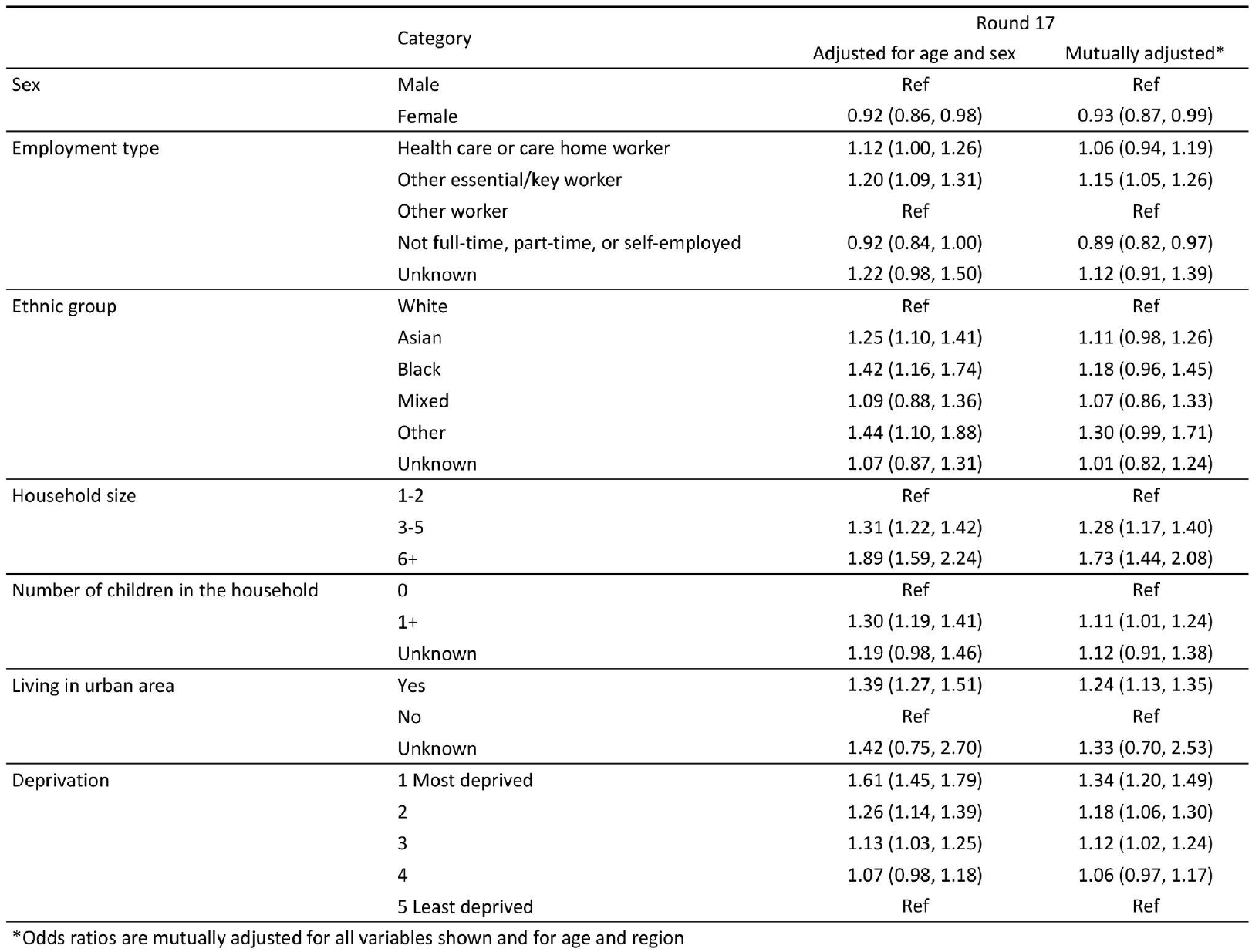
Multivariable logistic regression for SARS-CoV-2 swab-positivity in round 17. Results are presented as Odds Ratios (95% confidence interval) adjusted for age and sex and additionally, for region and all other variables (mutually adjusted OR).

|  |  | Round 17 |  |
| --- | --- | --- | --- |
|  | Category | Adjusted for age and sex | Mutually adjusted* |
| Sex | Male | Ref | Ref |
|  | Female | 0.92 (0.86, 0.98) | 0.93 (0.87, 0.99) |
| Employment type | Health care or care home worker | 1.12 (1.00, 1.26) | 1.06 (0.94, 1.19) |
|  | Other essential/key worker | 1.20 (1.09, 1.31) | 1.15 (1.05, 1.26) |
|  | Other worker | Ref | Ref |
|  | Not full-time, part-time, or self-employed | 0.92 (0.84, 1.00) | 0.89 (0.82, 0.97) |
|  | Unknown | 1.22 (0.98, 1.50) | 1.12 (0.91, 1.39) |
| Ethnic group | White | Ref | Ref |
|  | Asian | 1.25 (1.10, 1.41) | 1.11 (0.98, 1.26) |
|  | Black | 1.42 (1.16, 1.74) | 1.18 (0.96, 1.45) |
|  | Mixed | 1.09 (0.88, 1.36) | 1.07 (0.86, 1.33) |
|  | Other | 1.44 (1.10, 1.88) | 1.30 (0.99, 1.71) |
|  | Unknown | 1.07 (0.87, 1.31) | 1.01 (0.82, 1.24) |
| Household size | 1-2 | Ref | Ref |
|  | 3-5 | 1.31 (1.22, 1.42) | 1.28 (1.17, 1.40) |
|  | 6+ | 1.89 (1.59, 2.24) | 1.73 (1.44, 2.08) |
| Number of children in the household | 0 | Ref | Ref |
|  | 1+ | 1.30 (1.19, 1.41) | 1.11 (1.01, 1.24) |
|  | Unknown | 1.19 (0.98, 1.46) | 1.12 (0.91, 1.38) |
| Living in urban area | Yes | 1.39 (1.27, 1.51) | 1.24 (1.13, 1.35) |
|  | No | Ref | Ref |
|  | Unknown | 1.42 (0.75, 2.70) | 1.33 (0.70, 2.53) |
| Deprivation | 1 Most deprived | 1.61 (1.45, 1.79) | 1.34 (1.20, 1.49) |
|  | 2 | 1.26 (1.14, 1.39) | 1.18 (1.06, 1.30) |
|  | 3 | 1.13 (1.03, 1.25) | 1.12 (1.02, 1.24) |
|  | 4 | 1.07 (0.98, 1.18) | 1.06 (0.97, 1.17) |
|  | 5 Least deprived | Ref | Ref |

\*Odds ratios are mutually adjusted for all variables shown and for age and region

Comparing the prevalence of swab-positivity in REACT-1 to publicly available data on hospitalisations, with appropriate scaling and lag between the two curves, showed a close correspondence through round 8 (6 to 22 January 2021), and an apparent reduced risk of hospitalisation through rounds 9 to 11 (4 February to 3 May 2021), a coming together in rounds 12 and 13 (20 May to 12 July 2021) as Delta replaced Alpha, and a further period of reduced risk during rounds 14 to 15 (9 September to 5 November 2021), and finally a coming together in December 2021 as Omicron took off (Figure 3). Trends for deaths showed a marked and consistent reduction in risk compared to prevalence of swab-positivity throughout rounds 9 to 16 (4 February to 14 December 2021).

**Figure 3.**
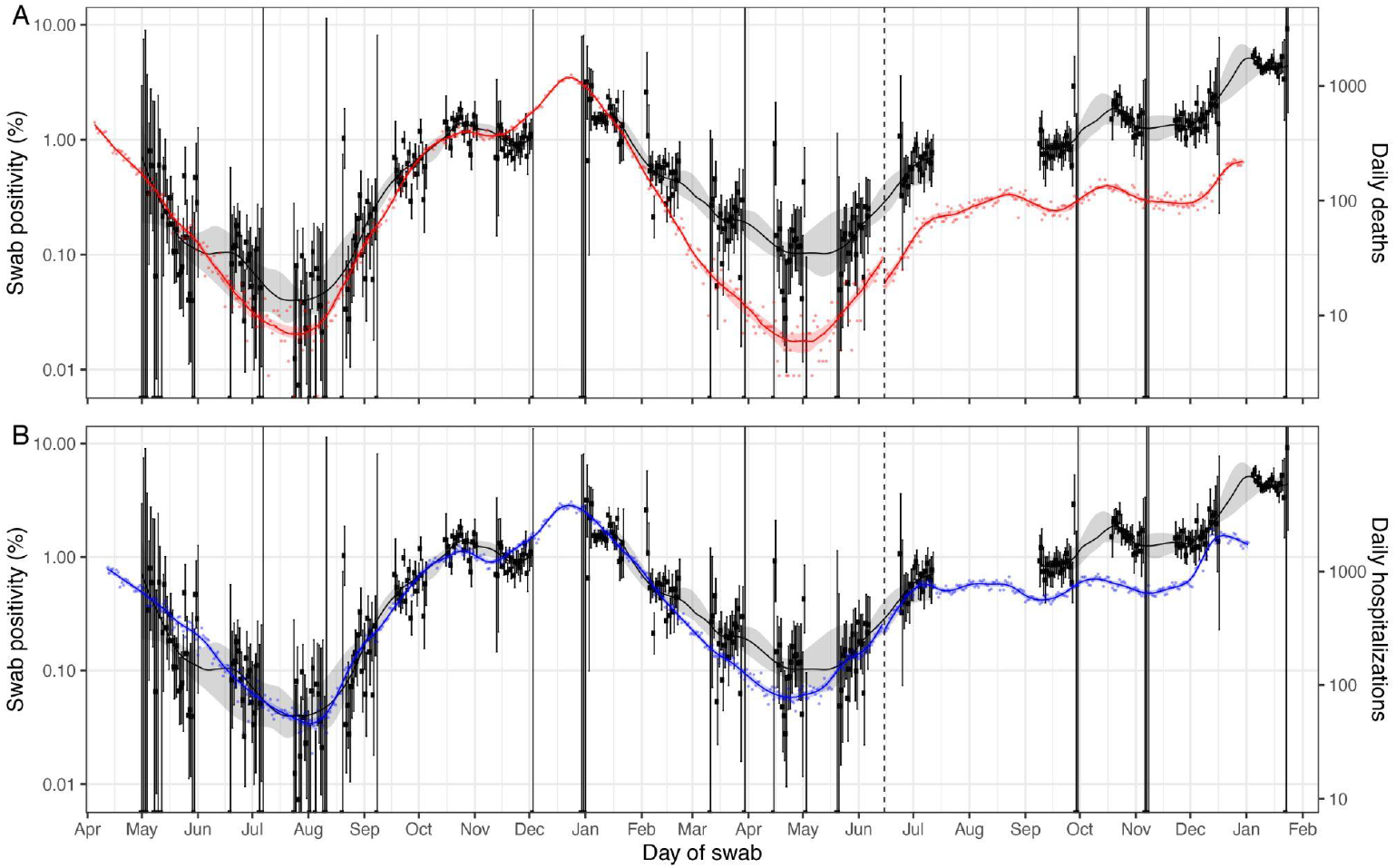
Comparison of COVID-19 daily deaths and hospitalisations with SARS-CoV-2 swab-positivity as measured in REACT-1. Daily swab-positivity for all 17 rounds of the REACT-1 study (black points with 95% confidence intervals, left-hand y-axis) with P-spline estimates for swab-positivity (solid black line, shaded area is 95% confidence interval). (A) Daily deaths in England (red points, right-hand y-axis) and P-spline model estimates for expected daily deaths in England (solid red line, shaded area is 95% confidence interval, right-hand y-axis). Daily deaths have been shifted by 25 days (95% CI, 25, 26) backwards in time along the x-axis up to 14 June 2021 and by 16 days (95% CI, 15, 16) thereafter. (B) Daily hospitalisations in England (blue points, right-hand y-axis) and P-spline model estimates for expected daily hospitalisations in England (solid blue line, shaded area is 95% confidence interval, right-hand y-axis). Daily hospitalisations have been shifted by 19 days (95% CI, 18, 20) backwards in time along the x-axis up to 14 June 2021 and by 16 days (95% 14, 18) thereafter. The scaling parameter estimates were 0.060 (0.058, 0.062) and 0.24 (0.23, 0.25) for deaths and hospitalisations, respectively.

## Discussion

The Omicron epidemic is further advanced in England than in many other countries. During December 2021 Omicron almost completely replaced Delta, with the peak in prevalence coming around six weeks after the first Omicron infection was identified in England. As of mid-January 2022, 0.80% of Omicron infections were BA.2 sublineage which has been designated a variant under investigation.^28^ Our data show an increase in the proportion of daily infections from BA.2 compared to BA.1 and its sublineage BA.1.1 with an R advantage of 0.46. We also detected one of the earliest instances of BA.3 in England ^29^. During round 17, we observed a drop in prevalence from the peak with a levelling off from mid-January, but still at extremely high levels. However, the dynamics underlying these population-level trends are complex with the prevalence falling in adults but rising in children through January 2022, likely the consequence of the peak occurring during the end-of-year school break, causing a delay to school-based transmission among children.

As a result of the rapid rise in Omicron infections, we saw the highest prevalence ever observed in the REACT-1 study, nearly three-fold higher than at the peak of the second wave in January 2021, with a near twelve-fold increase in the oldest age group (75 years and over) since December 2021. An estimated 20 to 40 times greater antibody titre is required for neutralisation of Omicron than for Delta,^30^ although individuals who received a third vaccine dose had increased neutralisation of the Omicron variant.^31,32^ Among infected individuals, in round 17, the distribution of Ct values was similar in adults who had received three compared to two doses of vaccine, suggesting that the booster did not affect viral load for Omicron. Crucially, booster doses are highly effective at reducing the risks of severe disease^3,33,34^ which is a key consideration in planning the public health response. Our comparison of infection prevalence data from REACT-1 and public data on hospitalisations and deaths indicate that although trends in these serious outcomes continue to track infections (albeit with an extended time lag) this is at a lower level than previously, before widespread rollout of the vaccination campaign in England. Nonetheless, it will be important to monitor hospitalisations and deaths closely over the coming weeks in view of the continued high levels of infection, including among the older population, and as restrictions are lifted in England and elsewhere.^35,36^

Our study has limitations. In round 17, 12.2% of the invited participants returned swabs producing valid RT-PCR test results, which is similar to what was observed in round 16 (response rate 12.1%). We use weights, calculated for each participant in each round, to adjust for differential response rates in calculating prevalence estimates, but these corrections may not fully eliminate all biases. Our results on reported previous COVID-19 are based on self-reported data. While it is uncertain what proportion of these are reinfections or recent infections picked up due to the sensitivity of PCR testing, among the swab-positive participants reporting previous COVID-19, 64.4% reported a date of most recent positive test within 30 days prior to swabbing, most likely due to residual infection. On the other hand, it is likely that some previous infections were under-reported, especially those occurring in the first wave when routine PCR testing was not readily available. Changes in the way the swab samples were transported and tested may have introduced small changes in results across rounds, although these should not have affected within-round trends.

In conclusion, we have documented a substantial and rapid rise in infections from early December 2021 through January 2022 as the Omicron variant took hold and almost completely replaced Delta in England. Although we have subsequently seen falls in prevalence in adults, prevalence remains very high. Among school-aged children there has been a rise in prevalence as they returned to school in January after the end-of-year break. Vaccination (including the booster campaign) remains the mainstay of the defence against SARS-CoV-2 given the high levels of protection against hospitalisations.^28,33,34^ However, further measures beyond vaccination may be required if the very high rates of Omicron infection persist, despite Omicron appearing to be intrinsically less likely to cause severe disease.^28,33,34^

## Data Availability

Access to REACT-1 individual-level data is restricted to protect participants' anonymity.
Summary statistics, descriptive tables, and code from the current REACT-1 study are available at https://github.com/mrc-ide/reactidd (doi 10.5281/zenodo.5574472). REACT-1 study materials are available for each round at https://www.imperial.ac.uk/medicine/research-and-impact/groups/react-study/react-1-study-materials/
Sequence read data are available without restriction from the European Nucleotide Archive at https://www.ebi.ac.uk/ena/browser/view/PRJEB37886, and consensus genome sequences are available from the Global initiative on sharing all influenza data (GISAID).

## Data availability

Access to REACT-1 individual-level data is restricted to protect participants’ anonymity. Summary statistics, descriptive tables, and code from the current REACT-1 study are available at https://github.com/mrc-ide/reactidd (doi 10.5281/zenodo.5574472). REACT-1 study materials are available for each round at https://www.imperial.ac.uk/medicine/research-and-impact/groups/react-study/react-1-study-materials/ Sequence read data are available without restriction from the European Nucleotide Archive at https://www.ebi.ac.uk/ena/browser/view/PRJEB37886, and consensus genome sequences are available from the Global initiative on sharing all influenza data (GISAID).

## Ethics

We obtained research ethics approval from the South Central-Berkshire B Research Ethics Committee (IRAS ID: 283787).

## Public involvement

A Public Advisory Panel provides input into the design, conduct, and dissemination of the REACT research program.

## Contributors

PE and CAD are corresponding authors. PE, MC-H and CAD conceived the study and the analytical plan. MC-H, BB, OE, HWang, DH, JJ, DT and CEW performed the statistical analyses. HWang, OE, DH, BB, and MW curated the data. JE, CA, PJD, DA, WB, GT, GC, HW, AD provided study oversight and results interpretation. AJP and AJT generated the sequencing data. AD and PE obtained funding. All authors revised the manuscript for important intellectual content and approved the submission of the manuscript. PE, MC-H, CAD had full access to the data and take responsibility for the integrity of the data and the accuracy of the data analysis and for the decision to submit for publication.

## Acknowledgement

PE is Director of the Medical Research Council (MRC) Centre for Environment and Health (MR/L01341X/1, MR/S019669/1). PE acknowledges support from Health Data Research UK (HDR UK); the National Institute for Health Research (NIHR) Imperial Biomedical Research Centre; NIHR Health Protection Research Units in Chemical and Radiation Threats and Hazards, and Environmental Exposures and Health; the British Heart Foundation Centre for Research Excellence at Imperial College London (RE/18/4/34215); and the UK Dementia Research Institute at Imperial College London (MC_PC_17114). AJP acknowledges the support of the Biotechnology and Biological Sciences Research Council (BB/R012504/1). HW acknowledges support from an NIHR Senior Investigator Award, the Wellcome Trust (205456/Z/16/Z), and the NIHR Applied Research Collaboration (ARC) North West London. MC-H and BB acknowledge support from Cancer Research UK, Population Research Committee Project grant ‘Mechanomics’ (grant No 22184 to MC-H). MC-H acknowledges support from the H2020-EXPANSE (Horizon 2020 grant No 874627) and H2020-LongITools (Horizon 2020 grant No 874739). JE is an NIHR academic clinical fellow in infectious diseases. GC is supported by an NIHR Professorship. CAD acknowledges support from the MRC Centre for Global Infectious Disease Analysis, the NIHR Health Protection Research Unit in Emerging and Zoonotic Infections and the NIHR-funded Vaccine Efficacy Evaluation for Priority Emerging Diseases (PR-OD-1017-20007).

We thank key collaborators on this work – Ipsos MORI: Kelly Beaver, Sam Clemens, Gary Welch, Nicholas Gilby, Kelly Ward, Galini Pantelidou and Kevin Pickering; Institute of Global Health Innovation at Imperial College London: Gianluca Fontana, Justine Alford; School of Public Health, Imperial College London: Eric Johnson, Rob Elliott, Graham Blakoe; Quadram Institute, Norwich, UK: Nabil-Fareed Alikhan; North West London Pathology and Public Health England (now UKHSA) for help in calibration of the laboratory analyses; Patient Experience Research Centre at Imperial College London and the REACT Public Advisory Panel; NHS Digital for access to the NHS register; the Department of Health and Social Care for logistic support.

## Supplementary material

**Supplementary Table 1.** Unweighted and weighted prevalence of SARS-CoV-2 swab-positivity from REACT-1 across rounds 1 to 17.

| Round | Tested swabs | Positive swabs | Unweighted prevalence (95% CI) | Weighted prevalence (95% CI) | First sample | Last sample |
| --- | --- | --- | --- | --- | --- | --- |
| 1 | 120,620 | 159 | 0.13% (0.11%, 0.15%) | 0.16% (0.13%, 0.19%) | 01/05/20 | 01/06/20 |
| 2 | 159,199 | 123 | 0.08% (0.07%, 0.09%) | 0.09% (0.07%, 0.11%) | 19/06/20 | 07/07/20 |
| 3 | 162,821 | 54 | 0.03% (0.03%, 0.04%) | 0.04% (0.03%, 0.05%) | 24/07/20 | 11/08/20 |
| 4 | 154,325 | 137 | 0.09% (0.08%, 0.11%) | 0.13% (0.01%, 0.15%) | 20/08/20 | 08/09/20 |
| 5 | 174,949 | 824 | 0.47% (0.44%, 0.50%) | 0.60% (0.55%, 0.71%) | 18/09/20 | 05/10/20 |
| 6 | 160,175 | 1,732 | 1.08% (1.03%, 1.13%) | 1.30% (1.21%, 1.39%) | 16/10/20 | 02/11/20 |
| 7 | 168,181 | 1,299 | 0.77% (0.73%, 0.82%) | 0.94% (0.87%, 1.01%) | 13/11/20 | 03/12/20 |
| 8 | 167,642 | 2,282 | 1.36% (1.31%, 1.42%) | 1.57% (1.49%, 1.66%) | 06/01/21 | 22/01/21 |
| 9 | 165,456 | 689 | 0.42% (0.39%, 0.45%) | 0.49% (0.44%, 0.55%) | 04/02/21 | 23/02/21 |
| 10 | 140,844 | 227 | 0.16% (0.14%, 0.18%) | 0.20% (0.17%, 0.23%) | 11/03/21 | 30/03/21 |
| 11 | 127,408 | 115 | 0.09% (0.07%, 0.11%) | 0.10% (0.08%, 0.13%) | 15/04/21 | 03/05/21 |
| 12* | 108,911 | 135 | 0.12% (0.10%, 0.15%) | 0.15% (0.12%, 0.18%) | 20/05/21 | 07/06/21 |
| 13 | 98,233 | 527 | 0.54% (0.49%, 0.58%) | 0.63% (0.57%, 0.69%) | 24/06/21 | 12/07/21 |
| 14** | 100,527 | 764 | 0.76% (0.71%, 0.82%) | 0.83% (0.76%, 0.89%) | 09/09/21 | 27/09/21 |
| 15*** | 100,112 | 1,399 | 1.40% (1.33%, 1.47%) | 1.57% (1.48%, 1.66%) | 19/10/21 | 05/11/21 |
| 16**** | 97,089 | 1,192 | 1.23% (1.16%, 1.30%) | 1.41% (1.33%, 1.51%) | 23/11/21 | 14/12/21 |
| 17† | 102,174 | 4,073 | 3.99% (3.87%, 4.11%) | 4.41% (4.25%, 4.56%) | 05/01/22 | 20/01/22 |

\* Sampling strategy changed for round 12 and subsequent rounds. Therefore unweighted prevalence is not directly comparable with previous rounds
\*\* Including N=509 samples from 28-30 September 2021. Sample handling changed in round 14. Therefore prevalence is not directly comparable with previous rounds
\*\*\* Including N=93 samples (all negatives) from 6-8 November 2021, and N=86 samples with no collection/arrival dates
\*\*\*\* Including N=661 samples (including 12 positives ) from 15-17 December 2021
† Including N=862 (including 36 positive) from 21 to 24 January 2022

**Supplementary Table 2.** Proportion of each of the N=2,393 SARS-CoV-2 lineage detected in positive samples with at least 50% genome coverage from round 17. Results are based on 3,028 positive sequenced samples.

| Sub-lineage | N (2,393) | Percentage |
| --- | --- | --- |
| Delta/ Delta sub-lineages |  |  |
| AY.34.1 | 1 | 0.04% ( 0.01% , 0.24% ) |
| AY.36 | 1 | 0.04% ( 0.01% , 0.24% ) |
| AY.4 | 7 | 0.29% ( 0.14% , 0.60% ) |
| AY.4.2.1 | 1 | 0.04% ( 0.01% , 0.24% ) |
| AY.4.2.2 | 2 | 0.08% ( 0.02% , 0.30% ) |
| AY.4.3 | 1 | 0.04% ( 0.01% , 0.24% ) |
| AY.5 | 1 | 0.04% ( 0.01% , 0.24% ) |
| AY.75 | 1 | 0.04% ( 0.01% , 0.24% ) |
| AY.98.1 | 1 | 0.04% ( 0.01% , 0.24% ) |
| B.1.1.174 | 1 | 0.04% ( 0.01% , 0.24% ) |
| B.1.617.2 | 2 | 0.08% ( 0.02% , 0.30% ) |
| Omicron/ Omicron sub-lineages |  |  |
| B.1.1.529 | 13 | 0.54% ( 0.32% , 0.93% ) |
| BA.1 | 1,822 | 76.14% ( 74.39% , 77.80% ) |
| BA.1.1 | 519 | 21.69% ( 20.08% , 23.38% ) |
| BA.2 | 19 | 0.79% ( 0.51% , 1.24% ) |
| BA.3 | 1 | 0.04% ( 0.01% , 0.24% ) |

**Supplementary Table 3A.** Weighted prevalence of SARS-CoV-2 swab-positivity in round 16 and round 17 by sex, age, region, urban/rural area, employment type, and ethnic group.

| Variable |  | Round 16 |  |  | Round 17 |  |  |
| --- | --- | --- | --- | --- | --- | --- | --- |
|  |  | Positive | Total | Weighted Prevalence | Positive | Total | Weighted Prevalence |
| Sex | Male | 564 | 43,938 | 1.49% (1.36%, 1.64%) | 1,858 | 45,031 | 4.52% (4.29%, 4.76%) |
|  | Female | 628 | 53,147 | 1.34% (1.23%, 1.46%) | 2,215 | 57,141 | 4.30% (4.11%, 4.50%) |
|  | Unknown | 0 | 4 | * | 0 | 2 | * |
| Age | 05-11 | 236 | 4,811 | 4.74% (4.15%, 5.40%) | 396 | 5,287 | 7.85% (7.10%, 8.69%) |
|  | 12-17 | 119 | 5,235 | 2.31% (1.91%, 2.80%) | 255 | 5,351 | 5.20% (4.57%, 5.92%) |
|  | 18-24 | 20 | 2,093 | 0.93% (0.57%, 1.51%) | 132 | 2,649 | 4.96% (4.11%, 5.96%) |
|  | 25-34 | 89 | 6,961 | 1.38% (1.10%, 1.74%) | 392 | 7,683 | 5.08% (4.57%, 5.65%) |
|  | 35-44 | 205 | 11,925 | 1.71% (1.48%, 1.98%) | 599 | 12,348 | 5.02% (4.62%, 5.46%) |
|  | 45-54 | 219 | 16,302 | 1.32% (1.15%, 1.52%) | 678 | 16,996 | 4.05% (3.74%, 4.38%) |
|  | 55-64 | 188 | 20,905 | 0.97% (0.84%, 1.13%) | 777 | 21,590 | 3.71% (3.45%, 3.99%) |
|  | 65-74 | 94 | 19,353 | 0.48% (0.39%, 0.59%) | 597 | 20,088 | 3.06% (2.82%, 3.33%) |
|  | 75+ | 22 | 9,504 | 0.21% (0.13%, 0.32%) | 247 | 10,182 | 2.46% (2.16%, 2.80%) |
| Region | South East | 228 | 16,908 | 1.54% (1.33%, 1.77%) | 545 | 18,287 | 3.23% (2.94%, 3.55%) |
|  | North East | 43 | 4,289 | 1.00% (0.70%, 1.42%) | 273 | 4,541 | 6.86% (5.99%, 7.84%) |
|  | North West | 107 | 11,352 | 1.08% (0.86%, 1.35%) | 589 | 11,867 | 5.36% (4.88%, 5.88%) |
|  | Yorkshire and The Humber | 96 | 9,261 | 1.32% (1.05%, 1.66%) | 488 | 9,736 | 5.53% (4.98%, 6.13%) |
|  | East Midlands | 125 | 8,584 | 1.73% (1.42%, 2.11%) | 352 | 9,045 | 4.15% (3.69%, 4.67%) |
|  | West Midlands | 89 | 9,715 | 1.03% (0.81%, 1.31%) | 476 | 10,297 | 5.20% (4.70%, 5.75%) |
|  | East of England | 121 | 11,437 | 1.22% (0.99%, 1.49%) | 350 | 11,938 | 3.42% (3.05%, 3.83%) |
|  | London | 239 | 14,819 | 1.84% (1.59%, 2.12%) | 682 | 14,849 | 4.89% (4.49%, 5.31%) |
|  | South West | 144 | 10,724 | 1.57% (1.30%, 1.90%) | 318 | 11,614 | 2.92% (2.58%, 3.30%) |
| Living in urban area | Yes | 956 | 75,982 | 1.43% (1.33%, 1.54%) | 3,402 | 79,566 | 4.72% (4.54%, 4.90%) |
|  | No | 233 | 20,951 | 1.35% (1.17%, 1.55%) | 661 | 22,405 | 3.14% (2.89%, 3.42%) |
|  | Unknown | 3 | 156 | 2.25% (0.70%, 6.98%) | 10 | 203 | 6.07% (2.99%, 11.92%) |
| Employment type | Health care or care home worker | 92 | 7,694 | 1.41% (1.13%, 1.76%) | 379 | 7,966 | 5.31% (4.75%, 5.94%) |
|  | Other essential/key worker | 247 | 13,427 | 1.96% (1.70%, 2.25%) | 722 | 14,170 | 5.39% (4.97%, 5.84%) |
|  | Other worker | 525 | 38,338 | 1.52% (1.38%, 1.68%) | 1,586 | 38,328 | 4.57% (4.32%, 4.83%) |
|  | Not full-time, part-time, or self-employed | 303 | 35,679 | 1.06% (0.92%, 1.21%) | 1,281 | 39,792 | 3.52% (3.30%, 3.75%) |
|  | Unknown | 25 | 1,951 | 1.19% (0.77%, 1.83%) | 105 | 1,918 | 5.70% (4.63%, 7.00%) |
| Ethnic group | White | 1,003 | 85,420 | 1.35% (1.26%, 1.45%) | 3,421 | 89,773 | 4.13% (3.98%, 4.28%) |
|  | Asian | 66 | 4,874 | 1.64% (1.24%, 2.17%) | 297 | 5,383 | 6.58% (5.76%, 7.50%) |
|  | Black | 38 | 1,833 | 2.10% (1.48%, 2.98%) | 105 | 1,730 | 6.55% (5.33%, 8.01%) |
|  | Mixed | 31 | 1,600 | 2.03% (1.40%, 2.94%) | 91 | 1,745 | 5.56% (4.48%, 6.88%) |
|  | Other | 16 | 984 | 1.81% (1.08%, 3.01%) | 58 | 963 | 7.35% (5.57%, 9.63%) |
|  | Unknown | 38 | 2,378 | 1.96% (1.40%, 2.74%) | 101 | 2,580 | 4.32% (3.48%, 5.34%) |

\* Prevalence estimates are not reported if based on less than 5 observations

**Supplementary Table 3B.**
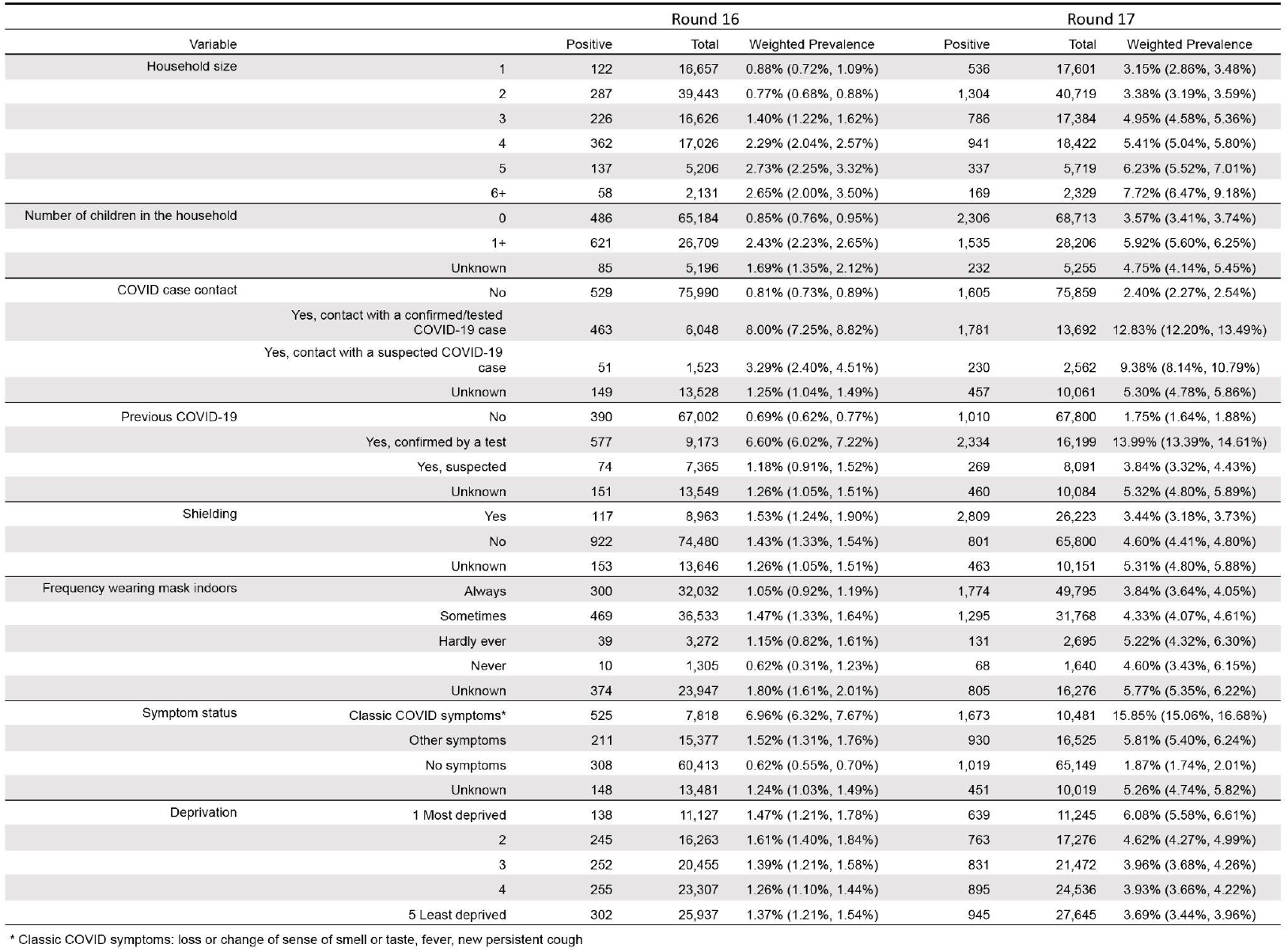
Weighted prevalence of SARS-CoV-2 swab-positivity in round 16 and round 17 by household size, number of children in the household, contact with a COVID-19 case, reported previous COVID-19, protective behaviours, symptom status and neighbourhood deprivation.

|  |  | Round 16 |  |  | Round 17 |  |  |
| --- | --- | --- | --- | --- | --- | --- | --- |
| Variable |  | Positive | Total | Weighted Prevalence | Positive | Total | Weighted Prevalence |
| Household size | 1 | 122 | 16,657 | 0.88% (0.72%, 1.09%) | 536 | 17,601 | 3.15% (2.86%, 3.48%) |
|  | 2 | 287 | 39,443 | 0.77% (0.68%, 0.88%) | 1,304 | 40,719 | 3.38% (3.19%, 3.59%) |
|  | 3 | 226 | 16,626 | 1.40% (1.22%, 1.62%) | 786 | 17,384 | 4.95% (4.58%, 5.36%) |
|  | 4 | 362 | 17,026 | 2.29% (2.04%, 2.57%) | 941 | 18,422 | 5.41% (5.04%, 5.80%) |
|  | 5 | 137 | 5,206 | 2.73% (2.25%, 3.32%) | 337 | 5,719 | 6.23% (5.52%, 7.01%) |
|  | 6+ | 58 | 2,131 | 2.65% (2.00%, 3.50%) | 169 | 2,329 | 7.72% (6.47%, 9.18%) |
| Number of children in the household | 0 | 486 | 65,184 | 0.85% (0.76%, 0.95%) | 2,306 | 68,713 | 3.57% (3.41%, 3.74%) |
|  | 1+ | 621 | 26,709 | 2.43% (2.23%, 2.65%) | 1,535 | 28,206 | 5.92% (5.60%, 6.25%) |
|  | Unknown | 85 | 5,196 | 1.69% (1.35%, 2.12%) | 232 | 5,255 | 4.75% (4.14%, 5.45%) |
| COVID case contact | No | 529 | 75,990 | 0.81% (0.73%, 0.89%) | 1,605 | 75,859 | 2.40% (2.27%, 2.54%) |
|  | Yes, contact with a confirmed/tested COVID-19 case | 463 | 6,048 | 8.00% (7.25%, 8.82%) | 1,781 | 13,692 | 12.83% (12.20%, 13.49%) |
|  | Yes, contact with a suspected COVID-19 case | 51 | 1,523 | 3.29% (2.40%, 4.51%) | 230 | 2,562 | 9.38% (8.14%, 10.79%) |
|  | Unknown | 149 | 13,528 | 1.25% (1.04%, 1.49%) | 457 | 10,061 | 5.30% (4.78%, 5.86%) |
| Previous COVID-19 | No | 390 | 67,002 | 0.69% (0.62%, 0.77%) | 1,010 | 67,800 | 1.75% (1.64%, 1.88%) |
|  | Yes, confirmed by a test | 577 | 9,173 | 6.60% (6.02%, 7.22%) | 2,334 | 16,199 | 13.99% (13.39%, 14.61%) |
|  | Yes, suspected | 74 | 7,365 | 1.18% (0.91%, 1.52%) | 269 | 8,091 | 3.84% (3.32%, 4.43%) |
|  | Unknown | 151 | 13,549 | 1.26% (1.05%, 1.51%) | 460 | 10,084 | 5.32% (4.80%, 5.89%) |
| Shielding | Yes | 117 | 8,963 | 1.53% (1.24%, 1.90%) | 2,809 | 26,223 | 3.44% (3.18%, 3.73%) |
|  | No | 922 | 74,480 | 1.43% (1.33%, 1.54%) | 801 | 65,800 | 4.60% (4.41%, 4.80%) |
|  | Unknown | 153 | 13,646 | 1.26% (1.05%, 1.51%) | 463 | 10,151 | 5.31% (4.80%, 5.88%) |
| Frequency wearing mask indoors | Always | 300 | 32,032 | 1.05% (0.92%, 1.19%) | 1,774 | 49,795 | 3.84% (3.64%, 4.05%) |
|  | Sometimes | 469 | 36,533 | 1.47% (1.33%, 1.64%) | 1,295 | 31,768 | 4.33% (4.07%, 4.61%) |
|  | Hardly ever | 39 | 3,272 | 1.15% (0.82%, 1.61%) | 131 | 2,695 | 5.22% (4.32%, 6.30%) |
|  | Never | 10 | 1,305 | 0.62% (0.31%, 1.23%) | 68 | 1,640 | 4.60% (3.43%, 6.15%) |
|  | Unknown | 374 | 23,947 | 1.80% (1.61%, 2.01%) | 805 | 16,276 | 5.77% (5.35%, 6.22%) |
| Symptom status | Classic COVID symptoms* | 525 | 7,818 | 6.96% (6.32%, 7.67%) | 1,673 | 10,481 | 15.85% (15.06%, 16.68%) |
|  | Other symptoms | 211 | 15,377 | 1.52% (1.31%, 1.76%) | 930 | 16,525 | 5.81% (5.40%, 6.24%) |
|  | No symptoms | 308 | 60,413 | 0.62% (0.55%, 0.70%) | 1,019 | 65,149 | 1.87% (1.74%, 2.01%) |
|  | Unknown | 148 | 13,481 | 1.24% (1.03%, 1.49%) | 451 | 10,019 | 5.26% (4.74%, 5.82%) |
| Deprivation | 1 Most deprived | 138 | 11,127 | 1.47% (1.21%, 1.78%) | 639 | 11,245 | 6.08% (5.58%, 6.61%) |
|  | 2 | 245 | 16,263 | 1.61% (1.40%, 1.84%) | 763 | 17,276 | 4.62% (4.27%, 4.99%) |
|  | 3 | 252 | 20,455 | 1.39% (1.21%, 1.58%) | 831 | 21,472 | 3.96% (3.68%, 4.26%) |
|  | 4 | 255 | 23,307 | 1.26% (1.10%, 1.44%) | 895 | 24,536 | 3.93% (3.66%, 4.22%) |
|  | 5 Least deprived | 302 | 25,937 | 1.37% (1.21%, 1.54%) | 945 | 27,645 | 3.69% (3.44%, 3.96%) |

\* Classic COVID symptoms: loss or change of sense of smell or taste, fever, new persistent cough

**Supplementary Table 4.** Distribution of self-reported dates of most recent positive SARS-CoV-2 test in REACT-1 participants from round 17. Results are presented for those reporting previous COVID-19 and those who did not, and for positive and negative swabs separately.

|  |  |  |  | Number of days prior to swabbing for most recent positive test |  |  |  |  |  |
| --- | --- | --- | --- | --- | --- | --- | --- | --- | --- |
|  |  |  |  | Total (%) | 1-14 days (%) | 15-30 days (%) | >30 days (%) | Invalid (%) | Missing (%) |
| Previous COVID-19 | Yes-confirmed | Total | 16,199 | 12,518 (77.28%) | 2,241 (13.83%) | 2199 (13.57%) | 7,875 (48.61%) | 203 (1.25%) | 3,681 (22.72%) |
|  |  | Positive Swabs | 2,334 | 1,771 (75.88%) | 1,337 (57.28%) | 167 (7.16%) | 114 (4.88%) | 153 (6.56%) | 563 (24.12%) |
|  |  | Negative Swab | 13,865 | 10,747 (77.51%) | 904 (6.52%) | 2,032 (14.66%) | 7,761 (55.98%) | 50 (0.36%) | 3,118 (22.49%) |
|  | No | Total | 67,800 | 2 (0.00%) | 0 (0.00%) | 0 (0.00%) | 1 (0.00%) | 1 (0.00%) | 67,799 (100.00%) |
|  |  | Positive Swabs | 1,010 | 1 (0.10%) | 0 (0.00%) | 0 (0.00%) | 0 (0.00%) | 1 (0.10%) | 1,009 (99.90%) |
|  |  | Negative Swab | 66,790 | 1 (0.00%) | 0 (0.00%) | 0 (0.00%) | 1 (0.00%) | 0 (0.00%) | 66,789 (100.00%) |

\* Including N=93 samples from 6-8 November 2021
\*\* Including N=661 samples from 15-17 December 2021
\*\*\* Including N=862 samples from 21-24 January 2022

**Supplementary Figure 1.**
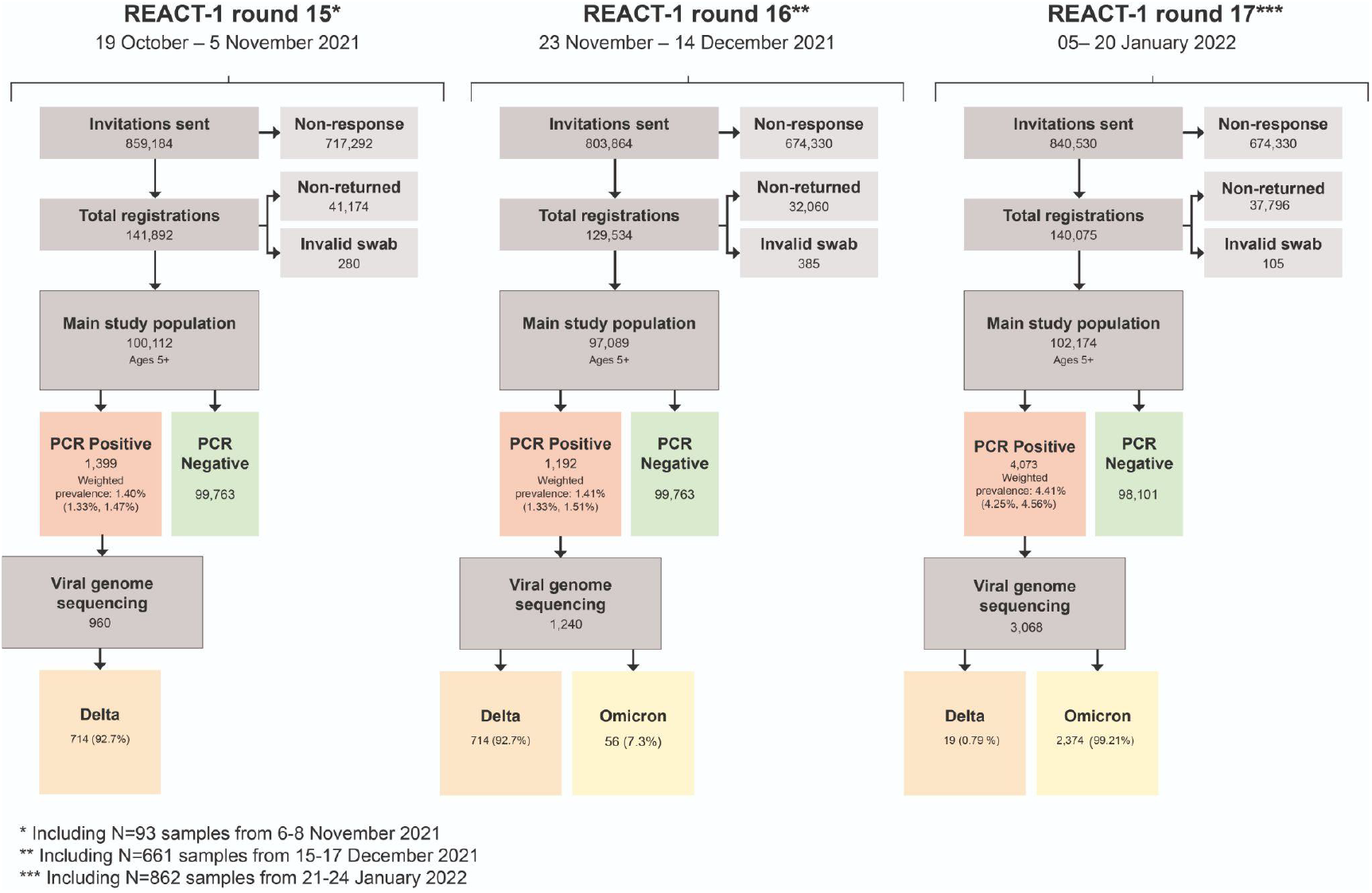
Flow chart showing numbers of participants in round 15 (19 October - 05 November 2021), round 16 (23 November - 14 December 2021) and round 17 (05 - 20 January 2022) of REACT-1.

**Supplementary Figure 2.**
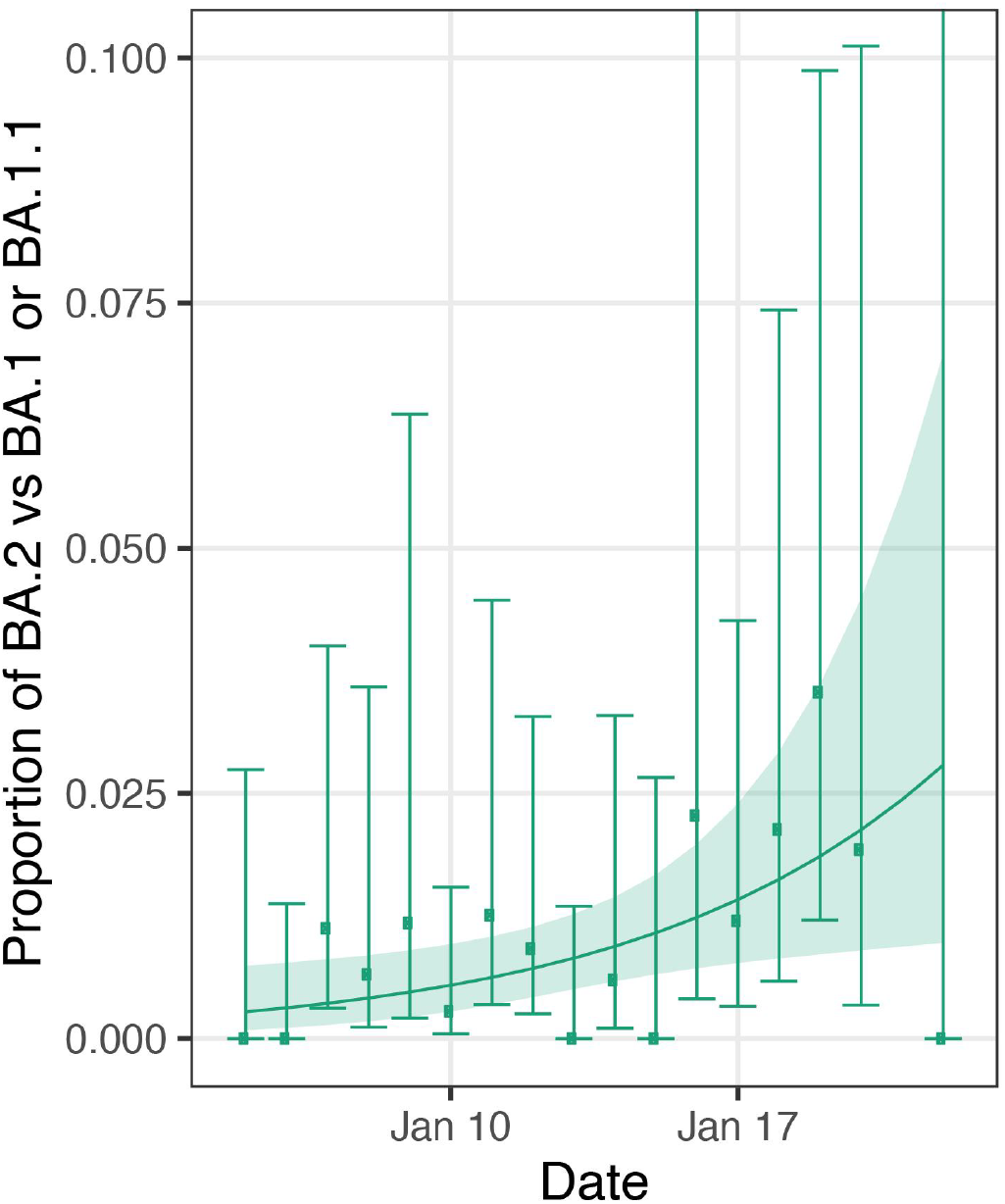
Daily proportion of BA.2 (vs BA.1 or BA.1.1) infections among positive swabs with determined lineage and at least 50% genome coverage in round 17. Point estimates are represented (dots) along with 95% confidence intervals (vertical lines). Smoothed estimates of the proportion are also shown (solid line) together with their 95% credible intervals (shaded regions). No sequencing data were available for 21 January 2022.

**Supplementary Figure 3.**
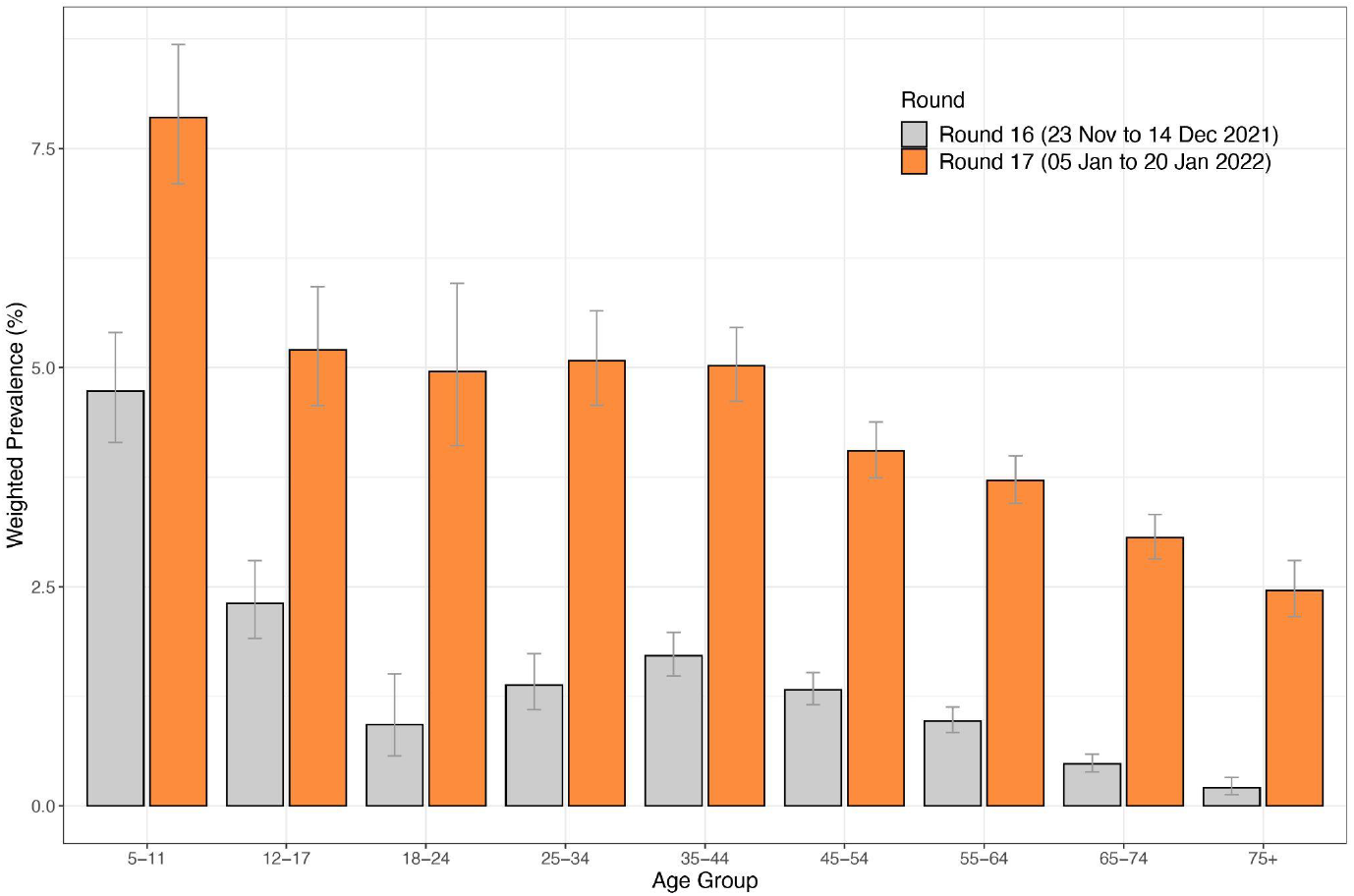
Weighted prevalence of SARS-CoV-2 swab-positivity by age group for round 16^5^ and round 17^6^. Bars show the weighted prevalence point estimates (grey for round 16 and orange for round 17), and the vertical lines represent the 95% confidence intervals.

**Supplementary Figure 4.**
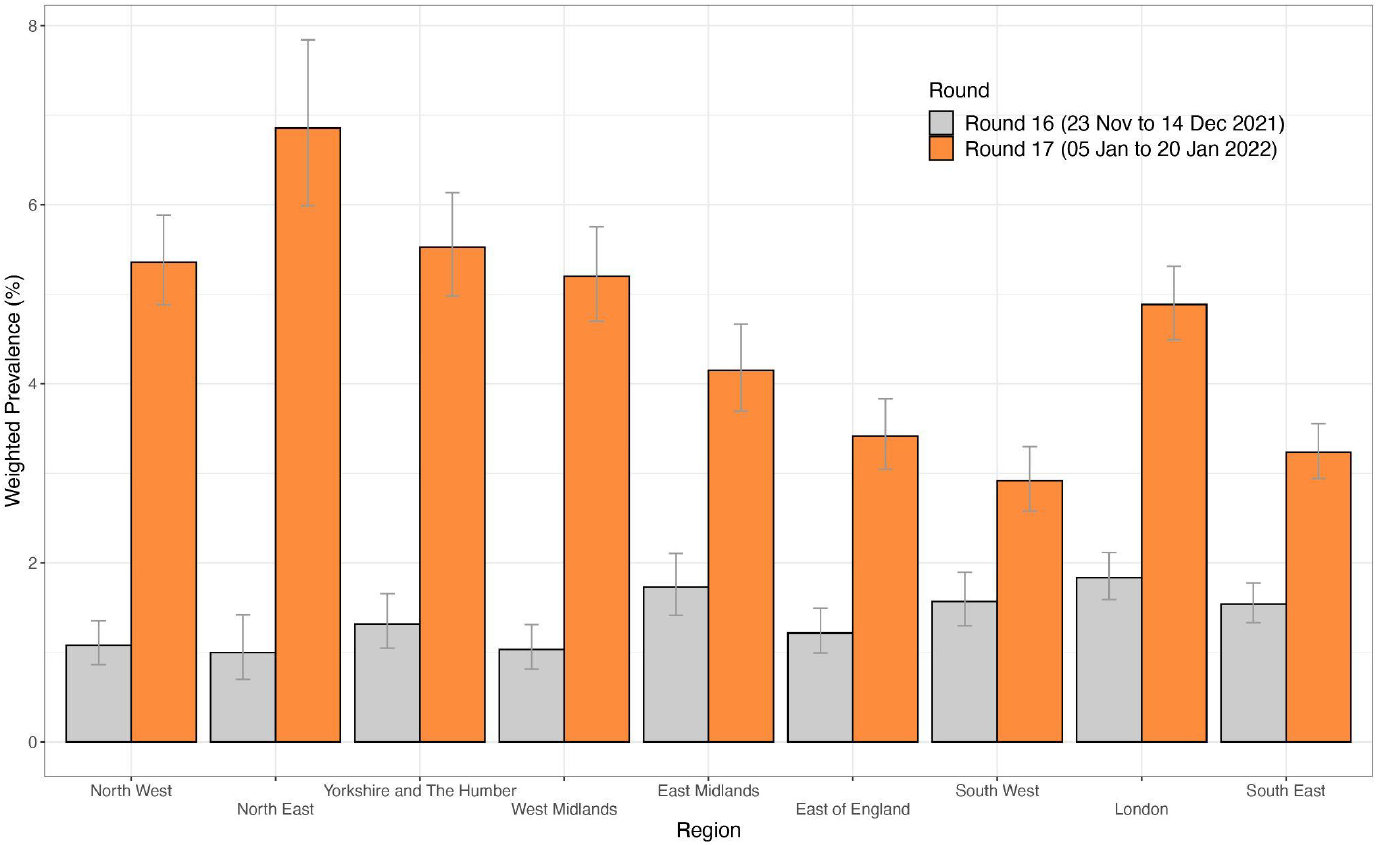
Supplementary Figure 4 Weighted prevalence of SARS-CoV-2 swab-positivity by region or round 16^7^ and round 17^8^. Bars show the weighted prevalence point estimates (grey for round 16 and orange for round 17), and the vertical lines represent the 95% confidence intervals.

**Supplementary Figure 5.**
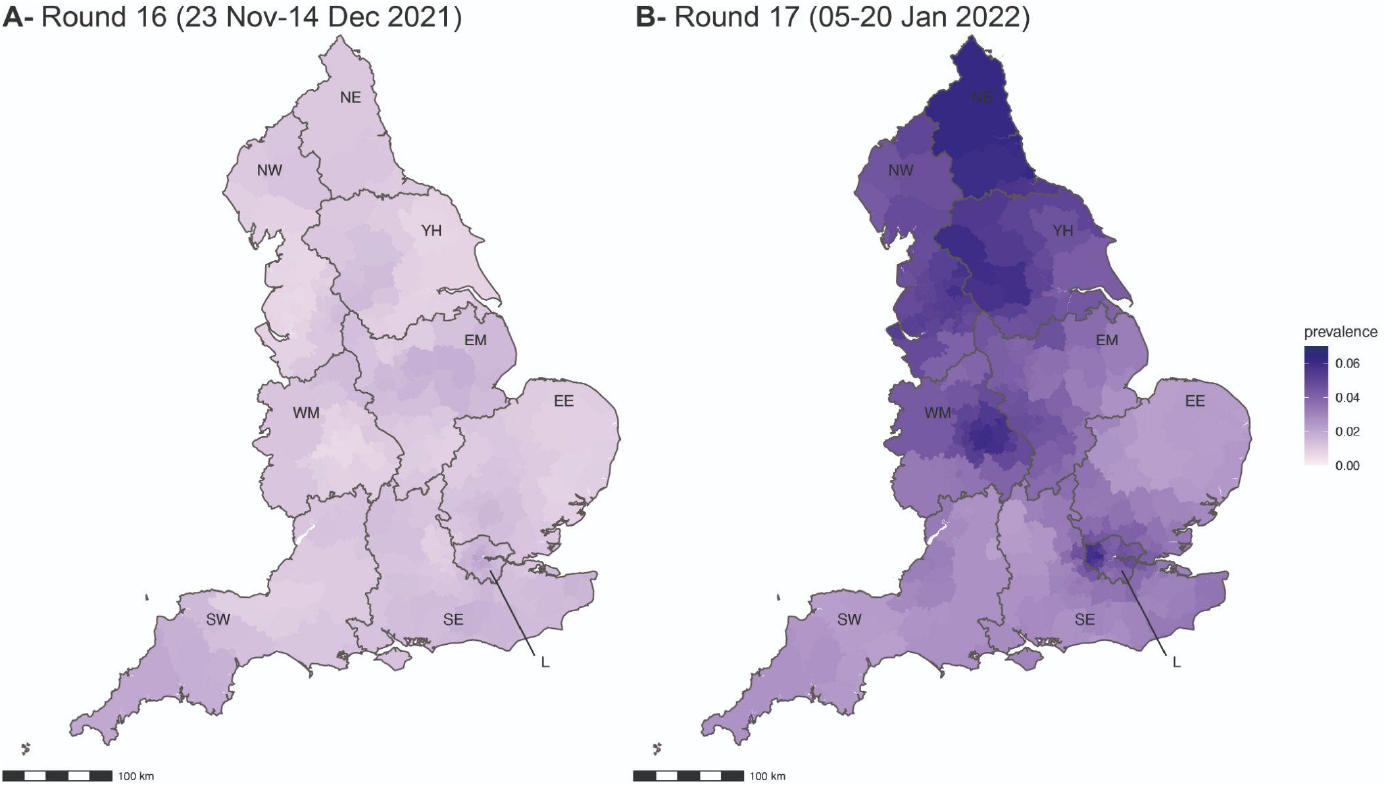
Neighbourhood smoothed average SARS-CoV-2 swab-positivity prevalence by lower-tier local authority area for round 16 (A) and round 17 (B). Neighbourhood prevalence calculated from nearest neighbours (the median number of neighbours within 30 km in the study). Average neighbourhood prevalence displayed for individual lower-tier local authorities for the whole of England. Regions: NE = North East, NW = North West, YH = Yorkshire and The Humber, EM = East Midlands, WM = West Midlands, EE = East of England, L = London, SE = South East, SW = South West

## Footnotes

1 Includes N=661 samples (12 positives) obtained from 15 to 17 December 2021.

2 Includes N=661 samples (12 positives) obtained from 15 to 17 December 2021.

3 Includes N=862 samples (36 positives) obtained from 21 to 24 January 2022

4 N=862 (including 36 positives) from 21-24 January 2022

5 Includes N=661 samples (12 positives) obtained from 15-17 December 2021

6 Includes N=862 samples (36 positives) from 21-24 January 2022

7 Includes N=661 samples (12 positives) obtained from 15-17 December 2021

8 Includes N=862 samples (36 positives) from 21-24 January 2022

